# Respiratory acclimatization and psychomotor performance after rapid ascent and during 3 weeks at 3100 m. A prospective cohort study in healthy

**DOI:** 10.1101/2024.11.18.24317470

**Authors:** Lara Muralt, Mona Lichtblau, Sayaka S Aeschbacher, Maya Bisang, Kay von Gruenigen, Talant M. Sooronbaev, Silvia Ulrich, Konrad E Bloch, Michael Furian

**Affiliations:** Pulmonary Division and Sleep Disorders Center, University Hospital of Zurich, Zurich, Switzerland; Research Department, Swiss University of Traditional Chinese Medicine, Bad Zurzach, Switzerland; National Center for Cardiology and Internal medicine, Bishkek, Kyrgyzstan; Swiss-Kyrgyz High Altitude Medicine and Research Initiative

## Abstract

**Background:** The acclimatization to high altitudes over several weeks is not extensively studied. Repeated physiological assessments were performed in healthy lowlanders staying at 3100m for 3 weeks. We hypothesized that over the course of 3 weeks, the arterial oxygen saturation (SpO_2_) improves and has favorable effects on psychomotor vigilance, postural control, and sleep.

**Methods:** 16 healthy volunteers (23-33y) underwent sleep studies during 1 night in Bishkek (760m), and during nights 1, 8 and 22 at Too-Ashu (3100m), Kyrgyzstan. The day after, reaction time (psychomotor vigilance test reaction time test, PVT) and postural control (center of gravity path length on balance board [COPL]) were assessed.

**Results:** Compared to 760m, mean nocturnal SpO_2_ dropped in the first night at 3100m from mean±SD 94.8±1.9% to 86.3±2.9% and recovered partially to 89.8±1.5% after 3 weeks (P<0.05 both comparisons to 760m). Corresponding median (quartiles) oxygen desaturation indices were 1.0/h(0.3;2.2), 6.5/h(4.5;12.1) and 6.4/h(4.2;11.1) time in bed (P<0.05 both comparisons to 760m). Median (quartiles) reaction times were 226ms(212;231), 236ms(210;259) and 228ms(212;246), P=NS, all comparisons. COPL worsened from 25.1±4.1cm to 27.1±4.1cm (P<0.05) and 26.4±3.7cm (P=NS compared to 760m).

**Conclusion:** In healthy lowlanders staying at 3100m, nocturnal SpO_2_ increased over 3 weeks after an initial drop but did not reach 760m baseline values. Postural control was impaired in the first week of acute exposure to high altitude despite improvements in hypoxemia. Altitude exposure had no effect on reaction time. Thus, acute and prolonged exposure to hypobaric hypoxia has differential effects on oxygenation, control of breathing, postural control and reaction time.

## INTRODUCTION

Many tourists, workers in mines or winter tourism, and researchers are exposed to high altitudes (>2500 m), while little is known about the adverse effects on health and performance during the acute and subacute acclimatization over several weeks. The few publications investigating a stay at altitude for several weeks have shown that the arterial oxygen saturation (SpO_2_) acutely decreases^1^ with altitude and redirects towards sea level values within weeks.^2-4^

During sleep at high altitudes, an oscillatory pattern of ventilation frequency and amplitude, known as periodic breathing, is commonly observed.^5,6^ However, it is uncertain if periodic breathing increases,^3^ decreases,^7^ or does not change^8^ during acclimatization. At altitudes between 3750 m and 6850 m, it has been observed that nocturnal periodic breathing increased during acclimatization over more than two weeks despite an improvement in SpO_2_. Whether a similarly prolonged ventilatory acclimatization takes place at altitudes of 3000 m to 4000 m, which has a higher relevance for most travelers and workers, remains open. Additionally, the acute exposure to hypoxia leads to impairments in postural control^9-13^ and cognitive performance.^4,14-18^ However, it is not known whether and to which extent postural control and cognitive performance are impaired due to the prolonged hypoxia and if they can recover during a few weeks of exposure to high altitude.

The current study aims at closing gaps in knowledge of nocturnal breathing, psychomotor performance, and postural control during acute exposure to 3100 m and subsequent acclimatization over three weeks. Understanding the underlying physiological acclimatization effects may help to prevent altitude induced accidents, mental and physical impairments, and to improve well-being at altitude.

We hypothesized that acute exposure to hypobaric hypoxia at 3100 m is associated with hypoxemia and could be associated with impaired cognitive performance and postural control, which could improve over the course of a three week long acclimatization period. Furthermore, we assumed that periodic breathing during sleep persist during the acclimatization period.

## METHODS

### Study design and setting

Baseline evaluations of this observational study were carried out in Bishkek (760 m), Kyrgyzstan. The assessments at high altitude were performed during the 1^st^, the 8^th^ and the 22^nd^ night and the following days (day 2, 9 and 23) after ascent to Too-Ashu, a high-altitude clinic located at 3100 m, 180 km south of Bishkek.

### Participants

Healthy lowlanders, 20 to 75 years of age, born, raised, and currently living below 1000 m were recruited. Participants with any disease that requires treatment, as well as heavy smokers (>20 cigarettes per day) were excluded. The study was approved by the Ethics Committee of the National Center of Cardiology and Internal Medicine, Bishkek, Kyrgyzstan (01-8/405) and all participants gave an informed written consent.

### Measurements

#### Sleep study

Respiratory polygraphy was performed from 23:00 to 06:00. Nasal pressure swings and pulse oximetry were measured using the ApneaLink™ device (ResMed Schweiz GmbH, Basel, Switzerland), which has been validated in several studies for screening of sleep disordered breathing.^19,20^ For the manual scoring the AASM Chicago Criteria were used for the definitions of apnea, hypopnea, and oxygen desaturation: apnea was defined as a ≥90% decrease in nasal pressure swings versus baseline for ≥10 s; hypopnea was defined as a decrease in nasal pressure swings by ≥50% to <90% for ≥10 s; oxygen desaturation was defined as a ≥4% decrease in SpO_2_.^21^ The apnea-hypopnea index and the oxygen desaturation index (ODI) were computed as number of events per hour of time in bed.

### Postural control

Balance tests were performed on a Wii Balance Board™ (WBB), 30 × 50 cm in size (Wii Balance Board, Redmond, WA, USA). The WBB has four pressure sensors with a sample rate of 100 Hz to measure the center of pressure (COP). The longer the path (cm) of the COP is, the more the person swayed (see Figure 1). Besides the COP path length (COPL), the maximal amplitude, and the sway velocity were measured in the anteroposterior and mediolateral axis.

**Figure 1:**
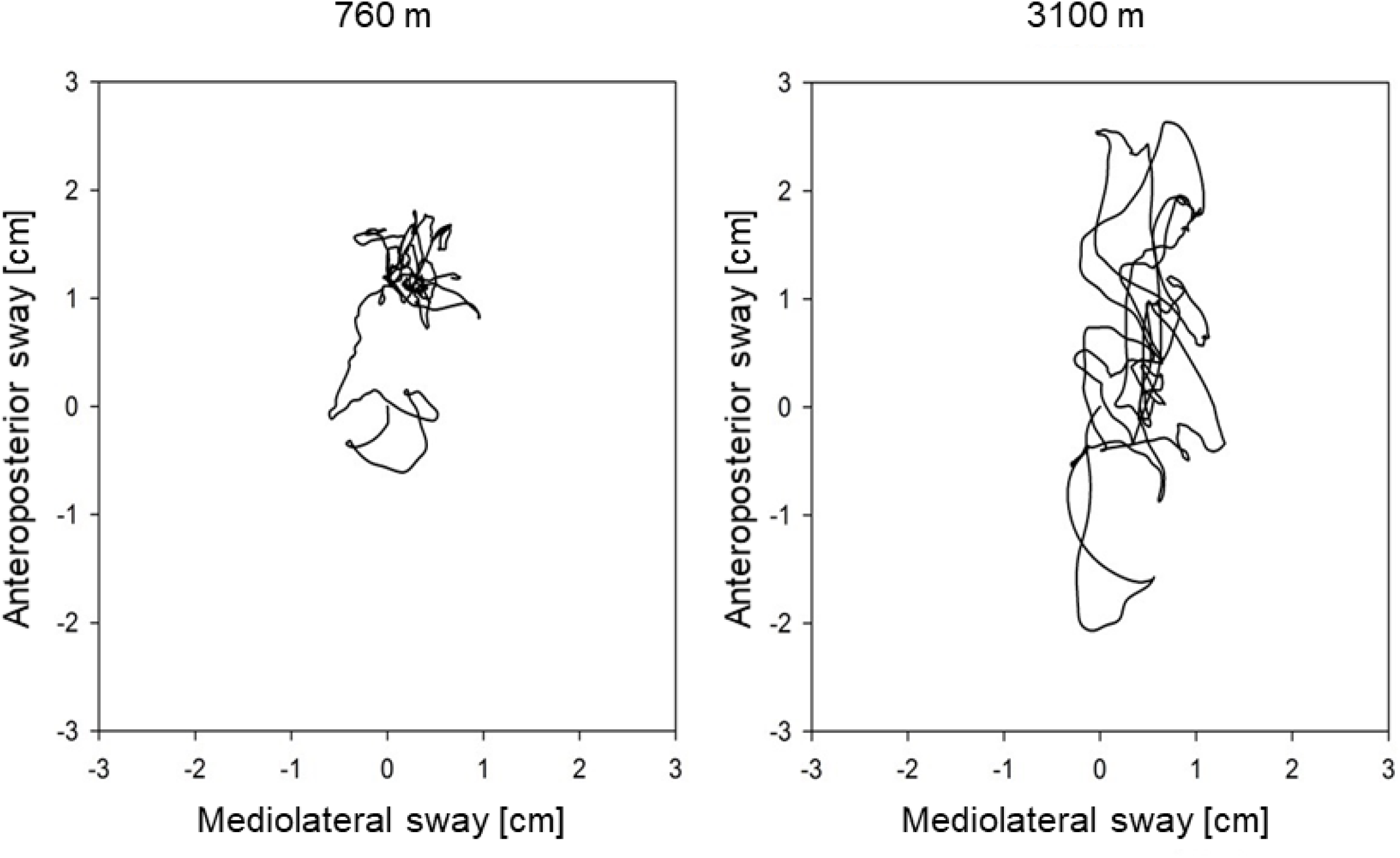
Example of a center of pressure path in an individual at 760 m and 3100 m.

The balance board was positioned 1.5 m in front of a wall. The participants stood on both legs with eyes open and feet in a 30° angle, as marked on the WBB. They were instructed to focus on a black dot on the wall at 1.7 m height, keep their hands beside the body and remain as still as possible while performing tests of 30 sec. The tests have been repeated 5 times and subsequently averaged.

A customized software (Labview 8.5, National Instruments, Austin, TX, U.S.A.) was used to track the COP and to analyze the data.^22^ The WBB was calibrated at each location and then once a week by placing a variety of known weights at different positions on the WBB. The assessment of the postural control using the WBB measurements has been validated by comparison with a laboratory based force platform.^22^

### Psychomotor and cognitive performance

The 10-minute-long psychomotor vigilance test (PVT) is a sustained-attention, reaction timed task test that measures cognitive performance.^23^ Participants performed the PVT alone in a darkened room and were instructed to look at a little lamp on the device, which lighted up in irregular time intervals of 2 to 10 sec. As soon as the light appeared, the participants had to press a button as fast as possible.^23,24^ The trail making test A (TMT-A) that requires connecting 90 encircled numbers by a line on a sheet of paper was administered.^25^ Participants performed two TMT-A versions at each testing session in random order, of which subsequently the mean was calculated.

### Questionnaires and clinical examinations

In the mornings after the sleep studies, participants rated their subjective sleep quality on a visual analogue scale labeled at 0 mm with “very bad” and at 100 mm with “very good”. They had to indicate the time until falling asleep in minutes, as well as the number and duration of awakenings during the past night. Acute mountain sickness (AMS) was assessed by the Environmental Symptoms Questionnaire cerebral score (AMSc) and AMS was defined as an AMSc ≥0.7.^26^ Moreover, daytime SpO_2_ was assessed by finger pulse oximetry.

### Outcomes and sample size estimation

The primary outcome was the difference in the mean nocturnal SpO_2_ (nSpO_2_) between night 1 and night 7 at 3100 m measured by pulse oximetry. Secondary outcomes were additional changes in nSpO_2_ of night 22 compared to night 1 at 3100 m. Additionally, changes in postural control, in cognitive performance, and in sleep patterns due to the ascent from 760 m to 3100 m, and the subsequent 3 weeks of acclimatization, were assessed. The sample size calculation was based on paired comparisons and a minimal important change in nSpO_2_ of 3% between night 1 and night 7 at 3100 m with a SD of 3%.^27,28^ To achieve a power of 80% with a significance level of 0.05 and accounting for a dropout rate of 20%, the required sample size was estimated to be 12 participants.

### Data Analysis and Statistics

All participants with SpO_2_ measurements of more than three hours during the four testing nights were included in the analysis. For the statistical analysis the software STATA/SE 13.1 was used. Data was tested for normality by the Shapiro-Wilks-test. Depending on data distribution, the effect of altitude and time at altitude were evaluated using non-parametric Friedman analysis of variance (ANOVA) followed by Wilcoxon matched pairs tests, or for parametric data, repeated measures ANOVA followed by paired t-tests. Post-hoc tests were only performed when the ANOVA showed a significant result.

The influence of independent parameters like the time at altitude, nSpO_2_, age, sex, body height, or body mass index on the COPL and the ODI has been evaluated with multivariate regression analysis. To obtain parametric data, the ODI had to be logarithmically transformed. The trial was registered under www.clinicaltrials.gov, NCT02451020.

## RESULTS

### Subjects

A total of 20 participants were invited. From those, three were excluded in the beginning of the study due to lack of time and one lost the nasal cannula during several nights and was therefore not included in the nocturnal respiratory analysis. Thus, nine men and seven women were included into the analysis (Table 1). One person fulfilled the AMS criteria on the second day at altitude with an AMSc of 0.733.

**Table 1.** Patient characteristics at 760 m.

|  |  |
| --- | --- |
| Number (% female) | 16 (43.8%) |
| Nationality, Kyrgyz / Swiss | 7 / 9 |
| Age, years | 27 ± 3 |
| Body weight, kg | 71.4 ± 9.6 |
| Body height, cm | 173.8 ± 8.5 |
| Body Mass Index, kg/m<sup>2</sup> | 23.7 ± 3.2 |
| FEV<sub>1</sub>, % predicted | 97.8 ± 15.4 |
| Arterial oxygen saturation, % | 97.5 ± 0.7 |

Data is shown in absolute numbers or mean ± SD. FEV<sub>1</sub>, forced expiratory volume during the first second of expiration.

### Sleep studies

The results are presented in Table 2 and Figure 2. The median (quartiles) of the analyzed time in bed was 6.8h (6.0 ; 7.5) and did not differ between the nights. In the first night at 3100 m, the mean nSpO_2_ was decreased (94.8 ± 1.9% vs 86.3 ± 2.9%) and time of nSpO_2_ <90% was increased (0.0% vs 97.5% of time in bed) compared to baseline but both improved partially over three weeks at altitude (Figure 2, Panel A and C). nSpO_2_ recovered from the first night at 3100 m to the eighth night from 86.3% to 88.8% (2.9% increase) and from the eighth to the 22^nd^ night from 88.8% to 89.8% (1.1% increase). Time of nSpO_2_ <90% recovered from 97.5% to 89.0% (8.7% decrease) and from 89.0% to 61.0% (31.5% decrease) respectively. In contrast, the ODI remained unchanged for 3 weeks after initial increase (Figure 2, Panel D). The regression analysis showed a positive correlation of altitude and ODI, even when adjusting for age, sex, and body mass index (Table 3). Body mass index was a significant positive predictor for a higher ODI, whereas female sex was associated with a lower ODI. Nocturnal breathing frequency did not change during the altitude sojourn, while the mean of nocturnal heart frequency increased significantly at altitude and remained elevated for at least one week.

**Table 2.** Effect of altitude and acclimatization on nocturnal breathing and daytime performance.

|  | **760 m** | **3100 m (night 1 /day 2)** | **3100 m (night 8 /day 9)** | **3100 m (night 22 /day 23)** | **P-Value\*** |
| --- | --- | --- | --- | --- | --- |
| Time in bed, hours | 6.7 (5.4 ; 6.9) | 7.1 (6.1 ; 7.8) | 6.8 (6.3 ; 7.3) | 7.3 (6.3 ; 7.7) | 0.466 |
| Nocturnal SpO<sub>2</sub>, % | 94.8 ± 1.9 | 86.3 ± 2.9 § | 88.8 ± 1.9 §, ¶ | 89.8 ± 1.5 §, ¶, ∞ | <0.001 |
| Oxygen Desaturation Index, events / hour | 1.0 (0.3 ; 2.2) | 6.5 (4.5 ; 12.1) § | 5.7 (3.6 ; 8.7) §, ¶ | 6.4 (4.2 ; 11.1) § | <0.001 |
| Sleep time < 90% SpO<sub>2</sub>, % of time in bed | 0.0 (0 ; 4) | 97.5 (87.0 ; 100.0) § | 89.0 (52.0 ; 96.5) § | 61.0 (41.5 ; 94.0) §, ¶ | <0.001 |
| Sleep time < 85% SpO<sub>2</sub>, % of time in bed | 0.0 (0.0 ; 2.0) | 36.0 (3.5 ; 76.5) § | 1.0 (0.0 ; 7.5) ¶ | 1.0 (0.0 ; 1.5) ¶ | <0.001 |
| Nocturnal breath frequency, events / min | 17.0 ± 1.9 | 17.3 ± 1.7 | 16.9 ± 1.7 | 16.6 ± 2.1 | 0.316 |
| Apnea-Hypopnea Index, events / hour | 5.6 (2.3 ; 7.8) | 6.4 (3.5 ; 8.4) | 6.2 (2.5 ; 8.6) | 9.1 (2.3 ; 18.6) | 0.249 |
| Heart frequency, bpm | 62 ± 6 | 68 ± 9 § | 70 ± 11 § | 66 ± 13 | 0.0032 |
| Awakenings, events | 1 (0 ; 2) | 3 (1 ; 3) § | 1 (1 ; 2) ¶ | 1 (0 ; 3) ¶ | 0.034 |
| Duration of awakenings, min | 6 (0 ; 13) | 11 (0 ; 50) § | 5 (1 ; 10) | 5 (0 ; 7) ¶ | 0.04 |
| Subjective sleep quality,<sup>1</sup> % | 70 (48 ; 77) | 55 (27 ; 79) | 73 (46 ; 84) | 63 (45 ; 81) | 0.460 |
| Daytime SpO<sub>2</sub>, % | 97.5 ± 0.7 | 94.0 ± 1.0 § | 93.9 ± 1.7 § | 94.2 ± 1.4 § | <0.001 |
| Median psychomotor vigilance test reaction time (PVT), ms | 226 (212 ; 231) | 236 (210 ; 259) | 237 (212 ; 244) | 228 (212 ; 246) | 0.115 |
| Trail making Test, s | 55 (50 ; 61) | 53 (45 ; 56) § | 49 (40 ; 54) §, ¶ | 47 (42 ; 53) §, ¶ | <0.001 |
| Center of pressure path length, cm | 25.1 ± 4.1 | 27.1 ± 4.1 § | 27.5 ± 3.2 § | 26.4 ± 3.7 | 0.048 |
| Anteroposterior amplitude, cm | 1.85 (1.46 ; 2.22) | 1.82 (1.67 ; 2.13) | 1.91 (1.62 ; 2.17) | 1.63 (1.47 ; 2.33) | 0.299 |
| Anteroposterior velocity, cm/s | 0.56 ± 0.11 | 0.64 ± 0.11 | 0.67 ± 0.09 § | 0.63 ± 0.14 | 0.027 |
| Mediolateral amplitude, cm | 1.36 (1.21 ; 1.56) | 1.31 (1.21 ; 1.43) | 1.32 (1.07 ; 1.50) | 1.22 (1.04 ; 1.44) | 0.131 |
| Mediolateral velocity, cm/s | 0.47 ± 0.10 | 0.51 ± 0.09 | 0.49 ± 0.10 | 0.48 ± 0.08 | 0.169 |

Nonparametric values are shown in median (interquartile range); parametric values in mean ± SD. <sup>1</sup>Subjective sleep quality was assessed by a 100-mm visual analogue scale, ranging from 0 “very bad” to 100 “very good”. \*Overall effects of repeated measures analysis of variance or Friedman ANOVA as appropriate; §P <0.05 vs. 760 m; ¶P <0.05 vs. night 1 or day 2 at 3100 m; ∞P <0.05 vs. night 8 or day 9 at 3100 m appraised with a paired t-test or Wilcoxon rank sum test as appropriate. SpO<sub>2</sub>, arterial oxygen saturation.

**Table 3.** Multivariate linear regression in Log10 oxygen desaturation index [events/hour].

|  | Coefficient | 95% CI | P-value |
| --- | --- | --- | --- |
| Time at altitude |  |  |  |
| 1<sup>st</sup> night at 3100 m (reference) |  |  |  |
| Night at 760 m vs 1<sup>st</sup> night at 3100 m | -2.14 | -2.73 to -1.55 | <0.001 |
| 8<sup>th</sup> night at 3100 m vs 1<sup>st</sup> night at 3100 m | -0.27 | -0.48 to -0.06 | 0.012 |
| 22<sup>th</sup> night at 3100 m vs 1<sup>st</sup> night at 3100 m | -0.24 | -0.64 to 0.16 | 0.238 |
| Age, years | 0.07 | -0.01 to 0.14 | 0.083 |
| Sex (female) | -0.56 | -1.06 to -0.06 | 0.028 |
| Body mass index, kg/m<sup>2</sup> | 0.10 | 0.06 to 0.14 | <0.001 |
| Intercept | -1.21 | -3.44 to 1.02 | 0.286 |

CI, confidence interval

**Figure 2:**
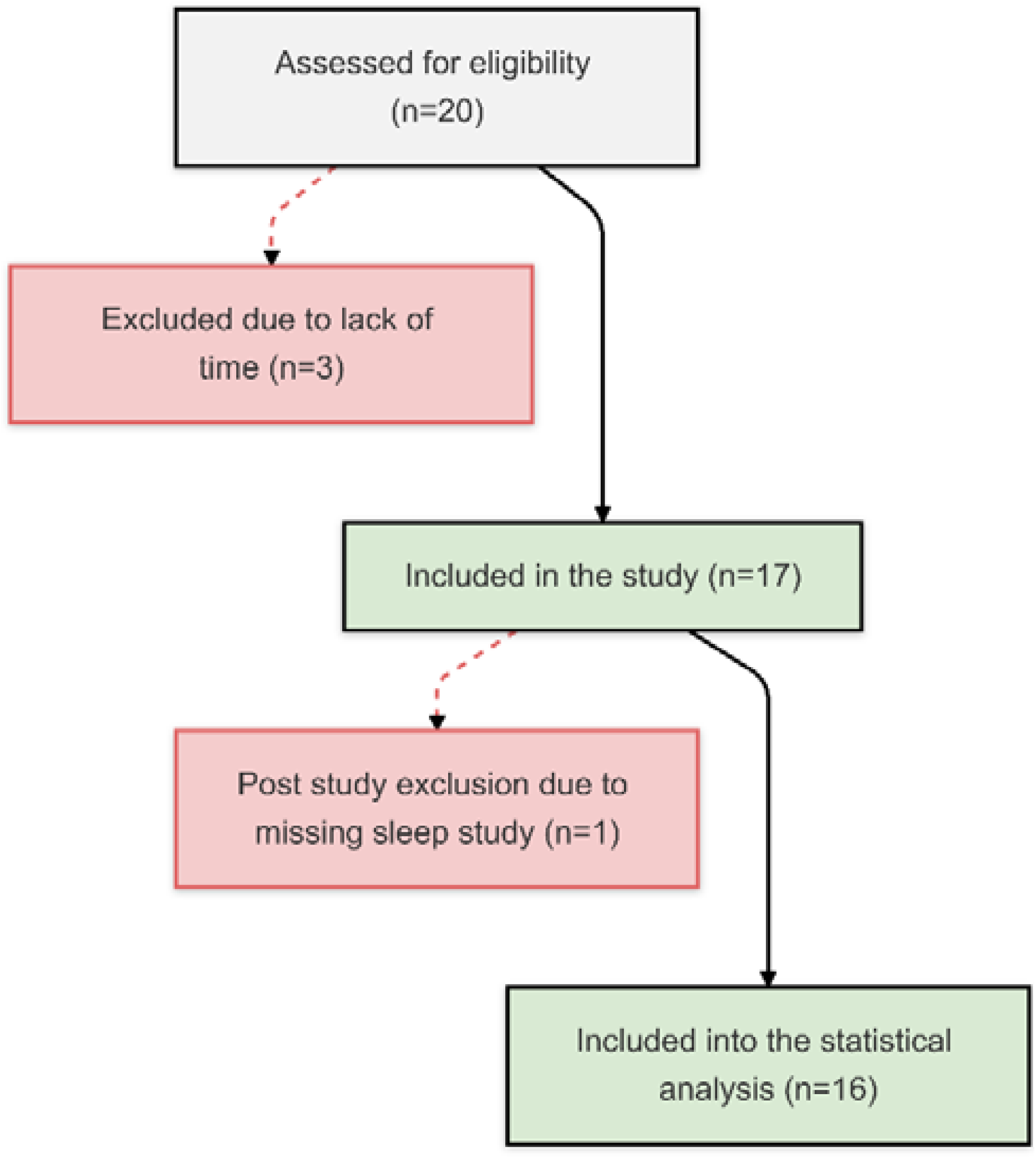
Study flow chart.

**Figure 3:**
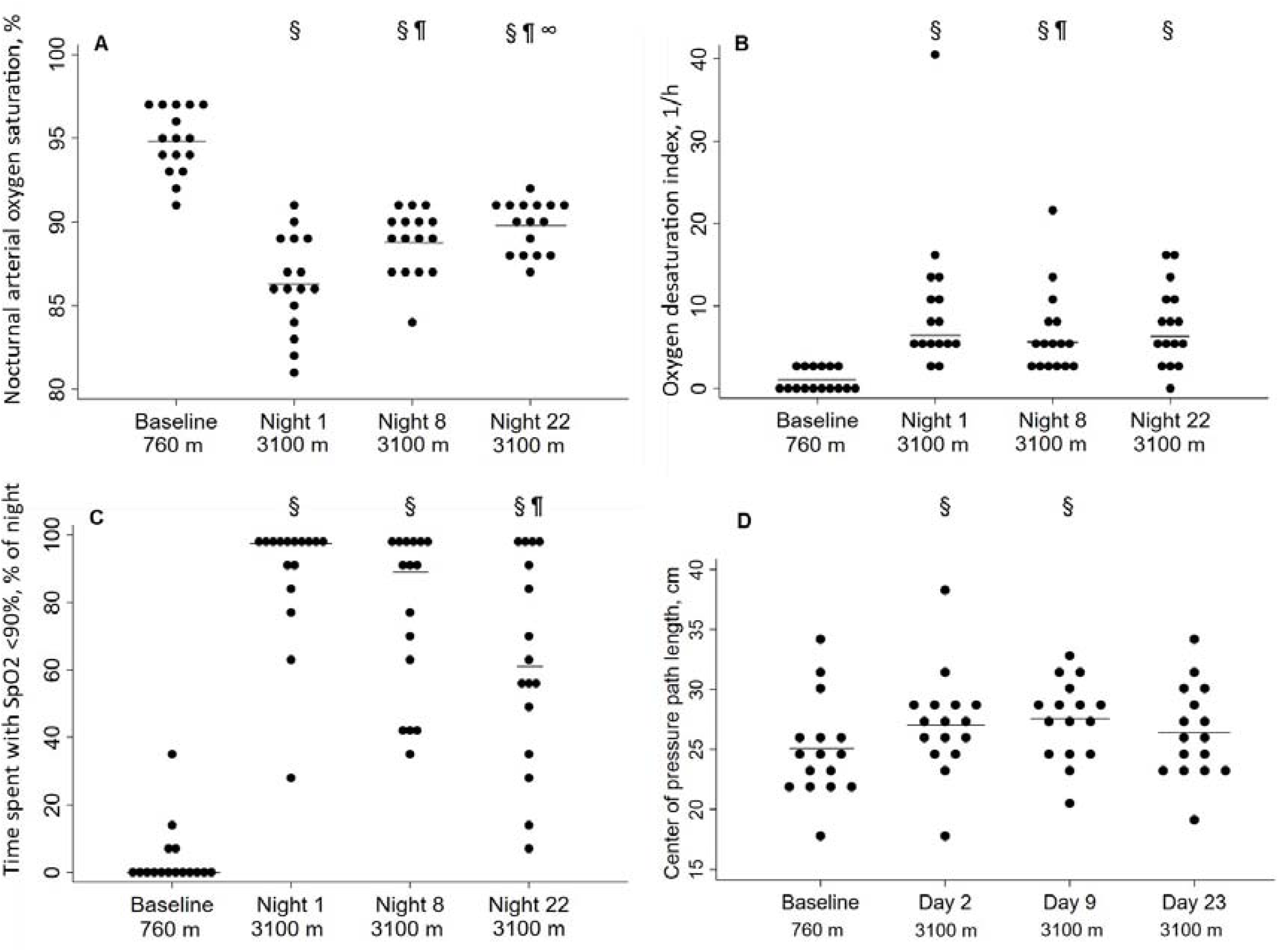
Effect of altitude and acclimatization. Panel A, nocturnal arterial oxygen saturation during the sleep [%]; Panel B, oxygen desaturation index [1/h]; Panel C, time spent with SpO_2_ <90% [% of time in bed]; Panel D, center of pressure path length [cm]. Each point represents a participant, the solid line represents the mean (Panel A and C) or the median (Panel B and D). ^§^P <0.05 vs. 760 m; ^¶^P <0.05 vs. night 1 at 3100 m; ^∞^P <0.05 vs. night 7 at 3100 m.

Subjective sleep quality at 3100 m remained unchanged compared to 760 m, but the participants reported more awakenings during the first night at 3100 m and estimated awakenings to be slightly longer.

### Postural control

With ascent from 760 m to 3100 m, COPL worsened significantly (Table 2, Figure 2 Panel B), remained impaired on day 9 but recovered after three weeks at 3100 m. The deterioration of postural control was mainly due to the change in the anteroposterior direction (Table 2). The regression analysis revealed a positive correlation of nSpO_2_ with altitude and negative correlation of nSpO_2_ on COPL during at least two weeks and no correlation of COPL with age, sex, or body height (Table 4).

**Table 4.** Multivariate linear regression in center of pressure path length [cm].

|  | **Coefficient** | **95% CI** | **P-value** |
| --- | --- | --- | --- |
| Daytime SpO<sub>2</sub>, % | -0.39 | -0.71 to -0.07 | 0.017 |
| Body height, cm | 0.06 | -0.14 to 0.26 | 0.543 |
| Sex (female) | -2.13 | -6.48 to 2.22 | 0.337 |
| Subjective sleep quality,<sup>1</sup> % | -0.02 | -0.06 to 0.02 | 0.290 |
| Intercept | 57.19 | 13.4 to 100.99 | 0.010 |

SpO<sub>2</sub> = arterial oxygen saturation. <sup>1</sup>Subjective sleep quality was assessed by a 100-mm visual analogue scale, ranging from 0 “very bad” to 100 “very good”.

### Cognition

There were no altitude dependent changes in the outcomes of PVT and TMT-A performance. However, the participants completed TMT-A within shorter time (i.e., had a better performance) at 3100 m vs. 760 m (Table 2).

## DISCUSSION

In this prospective cohort study, 16 healthy participants living below 1000 m have been exposed to 3100 m for three weeks. The sudden hypobaric hypoxia led to hypoxemia, which recovered partially over the course of the 3-week acclimatization period. Despite improvement in hypoxemia, the nocturnal periodic breathing pattern persisted for the entire three weeks. Moreover, postural control also remained impaired for over two weeks but normalized thereafter. No adverse altitude effect was observed in reaction time and cognitive performance. These findings suggest that acclimatization at 3100 m has differential effects on nocturnal oxygenation and control of breathing, postural control, and cognitive performance.

In a study in 51 healthy male lowlanders staying for 2 nights at 2590 m, the nSpO_2_ improved in the second night compared to the first night at 2590 m and the numbers of nocturnal desaturations redirected towards baseline values.^28^ Whereas in a study with 18 climbers at very high altitude (4497 to 6865 m), nSpO_2_ normalized over time while periodic breathing increased during two weeks.^3^ Additionally, in healthy individuals studied at an altitude of 4559 m periodic breathing increased further compared to the first night.^27^

In our study at 3100 m, ODI remained the same during the acclimatization period of 3 weeks. The varying tendency to periodic breathing during a prolonged stay at different altitudes may be related to changes in the sensitivity of the chemoreceptors to oxygen and carbon dioxide.^5,29^ An increase in ventilation may increase the partial pressure of oxygen but lower the partial pressure of carbon dioxide offsetting further hypoxic ventilatory stimulation. In our study, the breathing frequency remained unchanged with altitude, but information about the tidal volume was lacking.

Several authors showed the negative effect of simulated altitude on the postural control in healthy individuals through chamber experiments.^10,12,30^ Moreover, the postural control was impaired in 51 healthy men at moderate altitude of 1630 m.^9^ Several previous studies^12,13^ revealed a negative influence of acute hypobaric hypoxia on postural stability mainly in the anteroposterior direction, what we could confirm with this study. Presumably, hypoxia disturbs the fine interplay within the brain, neural afferents and motor output needed to maintain postural control. Indeed, in our multiple regression analysis the decreased daytime SpO_2_ was associated with an elongation of the COPL (Table 4), which has not been investigated intensively before.

Many travelers ascending from low altitude to high altitude report poor sleep quality during the first nights.^5^ However, subjective sleep quality was not reduced in this study, although the participants noted more and longer awakenings in the first night at 3100 m. Nonetheless, sleep parameters showed no correlation with postural control or cognitive performance, independent of daytime SpO_2_.

The literature on cognitive and psychomotor functions at high altitude reveals conflicting results – some reports indicating impairments especially at very high altitudes^4,31,32^ others no change.^28,33,34^ Reaction time remained unchanged with ascent to 3100 m and during the subsequent stay at high altitude. Possibly, a learning effect counteracting the adverse altitude effect on reaction time may account for these observations.

Little is known about sex differences during acute altitude exposure and acclimatization. We found that the female participants had less nocturnal oxygen desaturations, but otherwise we found no sex differences (Table 3). Unfortunately, information about any hormonal contraceptives and menstrual cycle, which might have influenced our results, was not available.

This study has several limitations. The small sample size may have attenuated other moderate effects in the acclimatization capacity when staying at 3100 m and potential sex differences could not be conclusively evaluated. Furthermore, depending on time constants, some acclimatization processes might have escaped the measurements after one and three weeks.

## Conclusions

In healthy lowlanders staying at 3100 m after rapid ascent, nSpO_2_ increased over three weeks but did not reach low altitude values. Postural control was impaired with acute exposure to altitude and remained impaired for more than one week, despite improvements in hypoxemia, suggesting slower reversibility. Vigilance and subjective sleep quality were not affected by altitude despite persistent nocturnal periodic breathing.

## Data Availability

All data produced in the present study are available upon reasonable request to the authors.

## Acknowledgments

Siemens Health Engineers provided some equipment for the study.

